# Mindfulness Coaching with Digital Lifestyle Monitoring Enhances Selective Attention in Medical Scientists

**DOI:** 10.1101/2024.01.04.24300716

**Authors:** Satish Jaiswal, Jason Nan, Suzanna R. Purpura, James K. Manchanda, Iris Garcia-pak, Dhakshin S. Ramanathan, Dawna Chuss, Deborah T. Rana, Ellen Beck, Paul A. Insel, Neil Chi, David M. Roth, Hemal H. Patel, Jyoti Mishra

## Abstract

Medical scientists have dual commitments to clinical care and research efforts. Such commitments can create hectic and stressful work schedules, which may impact on well-being and cognition. In this study, we tested the hypothesis that mindfulness coaching plus wearable-based lifestyle monitoring can benefit such individuals. We conducted a waitlist-controlled intervention study (n=43) with M.D./Ph.D. preclinical or graduate students and post M.D./Ph.D. trainees/faculty. Assessments of quantitative outcomes included subjective measures of burnout, mindfulness, self-compassion, and well-being, as well as objective cognitive assessments. The results showed no impacts on subjective measures. However, amongst objective cognitive measures, selective attention performance was significantly improved following the intervention (bias corrected effect size, Hedges’ g = 1.13, p=0.005). Extent of improvement in selective attention correlated with suppression of visual alpha oscillations – a neural marker for distractibility - measured using electroencephalography (EEG) (r=-0.32, p<0.05). Qualitative feedback showed that after receiving the intervention, participants in both groups equally rated the overall experience as “very good” (3.70 ±0.98 out 5). They also appreciated that the intervention emphasized healthy lifestyle behaviors, and contributed to mindfulness, compassion, and a sense of community. A majority (57%) of the participants reported that they expect to change their well-being related behaviors because of the intervention. Overall, this study suggests the utility of mindfulness coaching to improve selective attention skills in medical scientists and that more needs to be done to enhance subjective well-being in such individuals.

## 1. Introduction

The path of training to become a physician-scientist can be a long and potentially isolating journey. Such a career aims to bridge biomedical research and clinical care ^1^. It is a unique role to learn from patient experiences that inspire new research questions and to use scientific discovery to develop new diagnostics and therapeutics. The training in the U.S. often consists of pursuit of a dual-degree program, M.D./Ph.D., sometimes in a NIH-supported Medical Scientist Training Program (MSTP). Such programs transpire over 8-9 years before graduates undertake clinical residencies ^2^. Not only do these trainees toil through the rigorous workload of both medical school and graduate school, but their training integrates those environments and involves pressure to be efficient and accelerate through dual degree training. Transient states of belonging with peers can promote feelings of isolation among M.D./Ph.D. students, during post-M.D./Ph.D. training and for junior faculty ^3^. For M.D./Ph.D. trainees, transitions between medical school and graduate school can be particularly stressful and are challenging for M.D.s who undertake laboratory efforts ^4–7^. Physician-scientists in training may lack a supportive community and are at risk for poor well-being and burnout, a particular problem for individuals from groups underrepresented in the physician scientist community^8^. Physicians have a high rate of burnout, defined as “workplace stress that has not been successfully managed” (International Classification of Diseases, World Health Organization); such burnout rates are currently estimated at 44%, much higher than in other professions ^9,10^. The COVID19 pandemic exacerbated distress in healthcare workers and has been associated with dropout from training and reduction of work hours ^11,12^, which ultimately impacts on the entire healthcare system. Hence, a need exists for interventions that can help promote well-being, especially for individuals training as medical scientists ^13^.

Research has shown that group-based wellness interventions, such as group counseling and psychotherapy, improve well-being and cognition, likely benefitting from shared experiences that facilitate cohesion and interaction among the group members ^14–17^. In the past three decades, mindfulness-based interventions have received attention for alleviating stress and burnout symptoms. However, it can be challenging for physicians to accommodate their schedules to participate in traditional mindfulness trainings, which usually last up to 8 weeks, requiring 2.5 hours per week for didactics and practice ^18^. Also, having a fixed routine and coordinating in-person with a mindfulness coach can be a logistical challenge.

Given the need for implementing time-efficient well-being interventions for clinicians, particularly clinician scientists, we hypothesized that a digital intervention approach that integrated brief, online group mindfulness sessions led by expert coaches might be a useful way to enhance well-being. Thus, we provided each participant a smartwatch for monitoring and managing their stress and lifestyle. The intervention offered three coaching sessions over one academic quarter. In each session the coaches conducted focused mindfulness exercises and led an interactive discussion on health, well-being, self-compassion and resilience, and leadership training. The group intervention was scheduled to accommodate the preferred times of the program participants. We evaluated change on subjective scales of burnout, mindfulness, self-compassion and well-being and compared outcomes to a control group that did not receive the intervention. Such subjective measures have been used in other mindfulness-integrated intervention studies ^19–21^, and to test mindfulness programs in physician cohorts ^22–24^. Pflugeisen et al. ^22^ demonstrated post-intervention improvements in components of burnout, such as emotional exhaustion, as well as stress reduction, but that study lacked a control group. Schroeder et al. ^23^ did not find changes in subjective measures while Tickell et al. ^24^ showed improvement in depressive symptoms in the healthcare services but again without assessing a control group. Thus, the evidence for subjective benefits driven by mindfulness interventions in healthcare is weak; although self-compassion training in healthcare has shown benefit in a controlled study design ^25^ . Hence, here we also sought to measure objective benefits of the intervention, specifically whether it enhances cognitive functioning abilities and engenders cognitive neuroplasticity.

Cognitive abilities are fundamental to daily life functioning. We have implemented a scalable platform for measuring core aspects of cognition that delivers standard cognitive assessments synchronized with neural recordings using electroencephalography (EEG) ^26,27^. We have validated the reliability of this *BrainE©* platform and shown its utility in measuring cognitive changes across the lifespan ^28–32^. We also demonstrated its relevance to predict mental health ^33–36^. Here we deploy the *BrainE©* platform to measure post vs. pre-intervention changes in the well-being intervention group and compare outcomes with repeat assessments in a control group to control for practice effects. Specifically, we investigated whether the intervention can alter foundational abilities of selective attention, working memory, and interference processing of both non-emotional and emotional distractors. It is important to study different facets of cognition in this context as it has been postulated that mindfulness training may variably impact different forms of cognition depending on several factors, such as style of practice, target cohorts, coach characteristics, and environmental factors ^37^. Furthermore, EEG measures can provide insights into neural plasticity induced by the intervention ^38–42^, which is a novel approach in the context of interventions for the healthcare community.

## 2. Methods

### 2.1 Participants

A total of 43 physician-scientists participated in the study (mean age: 28.95± 4.36 years, range: 23-43 years, 42% female). Most (33 of 43 or 77%) participants were recruited from the University of California San Diego (UCSD)-MSTP; 10 of 43 (or 23%) had graduated from M.D./Ph.D. programs and were UCSD physicians/junior faculty at UCSD. Recruitment occurred via announcement at an annual retreat as well as by email. All participants provided written informed consent for study participation in accordance with the Declaration of Helsinki, and the UCSD Institutional Review Board approved all experimental procedures.

Data collection and intervention occurred from Fall 2021 through Summer 2022. Participants provided demographic information at the beginning of the study. Of the 43 participants, 22 were randomly assigned to the well-being intervention group (18 current MSTP trainees and 4 M.D./Ph.D. graduates) and 21 were assigned to a control group (15 current MSTP trainees and 6 M.D./Ph.D. graduates). The sample size within each group was powered to detect medium effect size pre/post differences (Cohen’s d >0.6) at a beta power of 0.8 and alpha level of 0.05. Between-group differences met criteria for investigating only large effect size outcomes (Cohen’s d >0.8) at a beta power of 0.8 and alpha level of 0.05. Effect sizes were calculated a priori using the G*Power software ^43^.

The study was registered in the International Standard Randomized Controlled Trial Number Registry ^44^. The well-being intervention group received the intervention during Fall-Winter quarter, while the control group received no intervention during this time. The control group eventually received the intervention during Spring-Summer quarter. All participants completed subjective and objective neuro-cognitive assessments before and after the first wave of the intervention, i.e., Fall-Winter quarter. At the end of Summer 2022, all participants in both groups completed a feedback survey.

No data were missing for subjective assessments. Two participants in the control group were missing cognitive and neural data. One participant in the well-being intervention group had corrupted neural data that could not be analyzed, hence, was marked as missing. Six participants (5 from the well-being intervention group and 1 from the control group) had missing data in the final qualitative survey.

### 2.2 Intervention

The intervention was a digital approach (delivered on Zoom) with group mindfulness peer mentoring sessions led by two psychological health faculty members as coaches. For these sessions, participants were divided into sub-groups by career stage: pre-clinical MSTP students, MSTP graduate students, or clinician/junior faculty. The coaches led three sessions (each 1.5 hours) spread over one academic quarter for each sub-group. The group meetings were held virtually to maximize accessibility and comfort for the participants and were scheduled per the participants’ mutually preferred times. In each session, the coaches conducted mindfulness exercises and led an interactive discussion on health, well-being, resilience, leadership training, and self-compassion. Participants were asked to turn on their videos for the duration of the sessions and to share personal opinions and life experiences. In these sessions, the primary goal for the coaches was to listen attentively and promote well-being. They also aimed to validate and acknowledge challenges that participants faced as physician-scientists in training or as early career faculty. Different mindfulness tools and perspectives were suggested and discussed by the facilitators and by the participants themselves to address work-life challenges. The participants were encouraged to share their sources of support (e.g., exploring sources of strength, stress management techniques, and spending time with loved ones). Brief self-compassion exercise was also offered and taught at each session such as soothing and supportive touch, and walking meditation modeled after Neff et al.^25^. The participants also discussed potential changes to the physician-scientist environment that would facilitate well-being, if implemented. At the end of each session participants were encouraged to have a tool or thought that was discussed and to prepare to share one at the next session. Intervention group participants were each provided a Garmin Vivosmart® 4 (a wearable device) to passively monitor and receive feedback on lifestyle factors (physical activity, heart rate, sleep quality and stress) via an app coupled to the Garmin for the duration of the intervention.

### 2.3 Assessments

These consisted of (1) subjective assessment scales, (2) objective cognitive and neural assessments and (3) a terminal feedback experience survey. Subjective and objective assessments were performed at pre- and post-intervention for the well-being intervention group while the control group performed these repeat assessments 3 months apart without any intervention and hence controlled for practice effects. The terminal feedback survey was administered after both groups had received the intervention.

**(1)** Subjective assessments were completed electronically and included validated behavioral self-report scales of mindfulness: 14-item mindful attention awareness scale (MAAS) ^45^, well-being: 7-item Short Warwick-Edinburgh mental well-being scale (SWEMWBS) ^46^, self-compassion: 12-item self-compassion scale ^47^, and burnout: 22-item Maslach Burnout Inventory (MBI) ^48,49^.
**(2)** Objective cognitive assessments were deployed on the *BrainE©* platform coded in Unity with simultaneous EEG ^26^. Participants visited the UCSD Neural Engineering and Translational Labs (NEAT Labs) for these assessments, which were delivered on a Windows-10 laptop. The Lab Streaming Layer (LSL) protocol was used to timestamp all stimuli and response events in all cognitive tasks ^50^. Each session lasted ∼40 minutes and consisted of four different cognitive tasks; each session additionally allowed for breaks between tasks to minimize fatigue. Tasks were run in the same order for all participants.

Figure 1 shows the stimulus sequence in each task. All four cognitive tasks had a standard trial structure of 500 ms central fixation “+” cue followed by a task-specific stimulus presented for task-specific duration and with a task-specific response window. This response window in each task was adaptive with a 3up-1down staircase scheme that maintains accuracy at ∼80% and engages the user by avoiding ceiling performance ^51,52^. An adaptive scheme also reduces practice effects that affect repeat assessment sessions. Further details of the adaptive scheme in each task are provided below.

**Figure 1.**
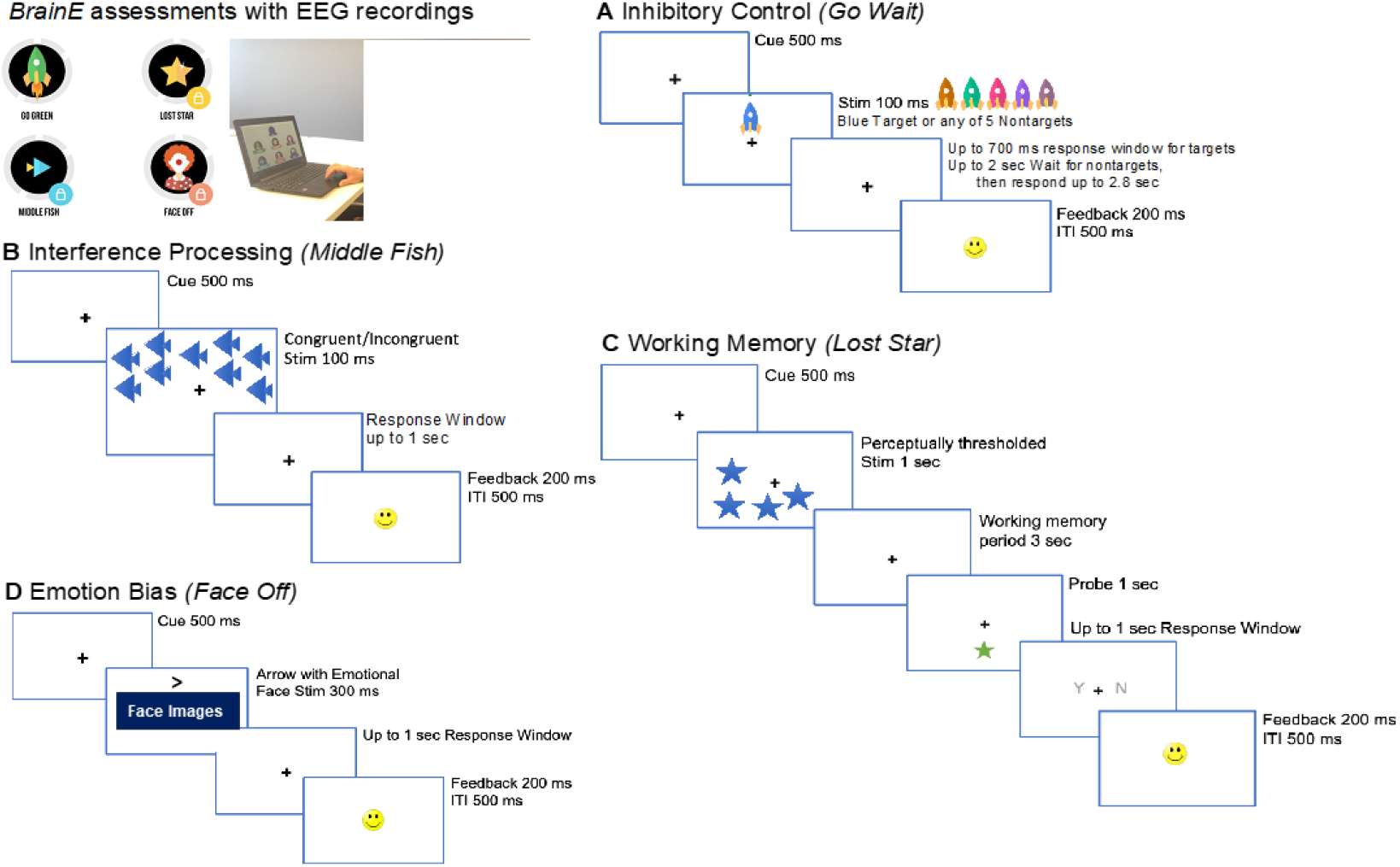
Cognitive assessments delivered on the *BrainE* platform. At top left, the *BrainE* assessment dashboard is shown with the wireless EEG recording setup. (**A**) Selective attention was measured on the ‘Go’ trials of the *Go Wait* task that required rapid and accurate responses to blue rocket targets. All other color rockets were non-targets on which responses were inhibited for up to 2 sec. (**B**) In the Flanker interference processing *Middle Fish* task, flanking fish may either face the same direction as the middle fish on congruent trials as shown, or the opposite direction on incongruent trials; participants were instructed to respond to the direction of the middle fish. (**C**) The visuo-spatial working memory task, *Lost Star*, was presented with perceptually thresholded stimuli and participants responded after a working memory period whether a probe star was positioned in one of the same locations as the prior test star stimuli. (**D**) The emotion bias task, *Face Off* presented neutral, happy, sad, or angry faces superimposed on an arrow, whose direction was discriminated by participants. The face images were obtained from the NimStim database, the readers should contact to the corresponding author to request those images.

Stimuli in each cognitive task were presented in a shuffled order across trials. Response in every task trial was followed by standard response feedback for accuracy as a smiley or sad face emoticon, presented 200 ms post-response for 200 ms duration, followed by a 500 ms inter-trial interval (ITI). ITI jitter within tasks was not applied to keep the task administration rapid. At the end of each task block, participants received a percent block accuracy score with a series of happy face emoticons (up to 10) to promote engagement.

**(2A)** <u>Selective Attention</u>: Participants accessed a game-like task, *Go Wait* modeled after the standard test of variables of attention ^53^. On each task trial, colored rockets were presented in the upper or lower central visual field. Participants were instructed to respond as rapidly as possible to blue colored rocket targets and wait to respond for 2 sec to distracting rockets of five other iso-luminant colors (shades of brown, teal, pink, purple). Iso-luminant colors were ensured using luminosity measurements in Photoshop. Luminance values for iso-luminant stimuli were 128, estimated as 0.30*R+0.59*G+0.11*B (where RGB are the Red/Green/Blue values of the chosen stimulus color). Post-fixation cue, a target/non-target stimulus appeared for 100 ms duration. For the blue rocket targets, the initial response window was set at 700 ms that adapted on each trial in a 3up-1down scheme, i.e., the response window reduced -33 ms after correct trials and increased +100 ms after incorrect trials. One happy face emoticon followed correct trials. If response was very rapid within 100-400 ms, then two happy face emoticons were presented for feedback to reinforce fast and accurate responding ^54^. For nontarget rockets, response times were not adaptive; participants waited for 2 sec at which time the fixation cue flashed briefly for 100 ms and then participants responded. Across two blocks, target and non-target trials were shuffled with 50% probability for 180 trials. Since task adaptivity was focused on target trials, processing speed on these trials was taken as the main selective attention outcome measure.

**(2B)** <u>Interference Processing</u>: Participants accessed the game-like task, *Middle Fish*, an adaptation of the Flanker assessment ^55–57^. Post-fixation on each trial, participants viewed an array of fish presented in the upper or lower central visual field for 100 ms. On each trial, participants had up to a 1 sec response window to detect the direction of the middle fish in the set (left or right) while ignoring the flanking distractor fish that were either congruent or incongruent to the middle fish, i.e., faced the same or opposite direction to the middle fish. Response windows were adapted on congruent trials in a 3up-1down scheme (−33 ms after correct trials and +100 ms after incorrect trials) and incongruent trial response windows matched that of the previous congruent trial. The fish flanking the middle fish interfere with the discrimination of the direction of the middle fish (left/right), hence, the task assesses interference processing. Task trials were shuffled with congruent/incongruent distractors in 1:1 ratio in 96 trials over two blocks. Processing speed on all trials was monitored as the main outcome.

**(2C)** <u>Working Memory</u>: Participants accessed a game-like task, *Lost Star*, which was based on the visuo-spatial Sternberg task ^58^. Post-fixation cue on each trial, participants viewed a spatially distributed test array of objects (i.e., a set of blue stars) for 1 sec. Participants were required to maintain the locations of these stars for a 3 sec delay period utilizing their working memory. A probe object (a single green star of 1 sec duration) was then presented in either the same spot as one of the original test stars, or in a different spot than any of the original test stars. The participant was instructed to respond whether the probe star had the same or different location as one of the test stars. 50% of task trials had the same probe star location as one of the test stars while 50% had different location, and presented in shuffled order. For each participant, we implemented this task at the threshold perceptual span, which was defined by the number of test star objects that the individual could correctly encode without any working memory delay. For this, a brief perceptual thresholding period preceded the main working memory task, allowing for equivalent perceptual load to be investigated across participants ^56^. During thresholding, the set size of test stars increased progressively from 1-8 stars based on accurate performance where 100% accuracy led to an increment in set size; <100% performance led to one 4-trial repeat of the same set size and any further inaccurate performance aborted the thresholding phase. The final set size at which 100% accuracy was obtained was designated as the individual’s perceptual threshold. Post-thresholding, the working memory task presented 48 trials over two blocks. Unlike the two previous tasks, this task was not speeded but instead adapted the working memory period in a 3up-1down scheme; if correct, the working memory period increased by +0.9 sec or if incorrect, it decreased by -0.3 sec. Both task item span and the length of the adapted working memory period were monitored as outcome measures.

**(2D)** <u>Emotion Bias</u>: Participants accessed the game-like assessment, *Face Off*, adapted from studies of attentional bias in emotional contexts ^59,60^. ^53^ The task integrated a standardized set of culturally diverse faces from the NimStim database ^61^. We used an equivalent number of male and female faces, each face with four sets of emotions: neutral, positive (happy), negative (sad) or threatening (angry), presented on equivalent number of trials in each task block. Post-fixation cue on each trial, participants viewed an emotional face with a superimposed arrow of 300 ms duration. The arrow occurred in either the upper or lower central visual field on equal number of trials. Participants responded to the direction of the arrow (left/right) within an ensuing 1 sec response window. For neutral emotion trials, this response window adapted in a 3up-1down scheme (−33 ms after correct trials and +100 ms after incorrect trials). All other emotion trials followed the same response window as their previous neutral emotion trial. This task evaluates emotion bias (or interference) as the emotional faces interfere with the discrimination of the direction (left/right) of the arrow on which they are superimposed. Participants completed 144 trials presented over three equipartitioned blocks. Processing speed across trials was monitored as the main outcome.

EEG data were collected in conjunction with all cognitive tasks using a 24-channel saline-soaked electrode cap with electrode locations as per the 10-20 system attached to a wireless SMARTING^TM^ amplifier. Signals were acquired at 500 Hz sampling frequency at 24-bit resolution. The Lab Streaming Layer (LSL) protocol was used to time-stamp EEG markers and integrate cognitive markers ^50^. Files were stored in xdf format.

**(3)** The end of year feedback survey was provided after all participants groups had undergone the intervention. The survey asked about participants’ overall experience on a 1 (poor) to 5 (excellent) Likert scale. It also asked participants to provide a Yes/No response as to whether they expected future behavior change due to the intervention. Participants also were asked what they particularly liked/learned from the program and/or what could be improved.

### 2.4 Data Analysis

Demographic characteristics were compared between the two groups using the non-parametric Wilcoxon rank sum test for age comparisons and *χ*^2^ (Chi-Square) test for gender and ethnicity comparisons.

#### 2.4.1 Behavioral and Cognitive

Normality of distributions was checked using the Levene’s test, and parametric or non-parametric repeated measure analysis of variance (rm-ANOVA) were conducted to interrogate group x session interactions. We applied the Greenhouse-Geisser correction to adjust for lack of sphericity. For any significant rm-ANOVA results, post-hoc analyses within each group, we used a paired t-test or the non-parametric equivalent Wilcoxon signed-rank test based on the normality of the distribution. For behavioral subjective assessments, we calculated scores on the mindfulness, well-being, self-compassion and burnout scales for both pre and post sessions.

The data for all cognitive tasks were analyzed for the task-relevant outcome measure. For the selective attention, interference processing and emotion bias tasks, processing speed was calculated at both pre and post sessions as log(1/RT), where RT is response time in seconds; thus, longer RTs have less speed while shorter RTs have higher processing speed. For the working memory task, item span and working memory delay period length were both outcomes. Outliers >3 median absolute deviation (mad) from median were removed and all metrics were verified for normal distributions prior to statistical analyses. Rm-ANOVAs were used to interrogate group x session interactions, and the Greenhouse-Geisser correction was applied to adjust for lack of sphericity. Additionally, false discovery rate (fdr) corrections were applied to account for multiple comparisons. Post-hoc analyses within each group used paired t-tests.

For the end of year feedback survey, overall experience ratings were compared between groups using Wilcoxon rank sum tests. Binary expected behavior change was analyzed using the Chi-Square test. Themes were extracted from qualitative responses and their proportional occurrence in each group were also compared using Chi-Square tests.

Effect sizes are reported for significant results as Cohen’s d, 0.2: small, 0.5: medium, 0.8: large ^62^.

#### 2.4.2 Neural Analysis

Neural data were analyzed for cognitive task(s) that showed any significant intervention-related cognitive performance effects. A uniform processing pipeline was applied to EEG data based on the LSL event markers. Data were pre-processed for computing event related spectral perturbations (ERSP) and for source localization. Data pre-processing utilized the EEGLAB toolbox in MATLAB ^63^. EEG data were first resampled at 250 Hz and filtered in the 1-45Hz range to exclude ultraslow DC drifts at <1HZ and high-frequency noise produced by muscle movements and external electrical sources at >45Hz. EEG data were average electrode referenced and epoched to stimulus onset based on the LSL timestamps, within the -1.0 to +1.0 sec event time window. The epoched data were then cleaned using the *autorej* function of EEGLAB, which automatically removes noisy trials (>5sd outliers rejected over max 8 iterations). EEG data were further cleaned by excluding signals estimated to be originating from non-brain sources, such as electro-oculographic, electromyographic or unknown sources, using the Sparse Bayesian learning (SBL) algorithm ^64,65^. This algorithm localizes EEG data similar to low-resolution electromagnetic tomography (LORETA) ^66^ with sparsity constraints applied to protect against false positives that are not biologically plausible. SBL yields source configurations from a few active regions, akin to independent component analysis, and are maximally independent from one another. Data from non-brain sources are removed to obtain clean subject-wise trial-averaged EEG. We verified that for cleaned data, channel peak activity in individual participant data did not exceed 3 mad from average channel activity across all subjects (total n = 40).

For ERSP calculations, we performed time-frequency decomposition of the epoched data using the continuous wavelet transform (cwt) function in MATLAB’s signal processing toolbox. Event-related synchronization (ERS) and event-related desynchronization (ERD) modulations were computed as baseline-corrected activity using the -750 ms to -550 ms time window prior to stimulus presentation as the baseline ^67^. Outliers >3 mad activity level were removed from the final ERSP matrices. The cleaned data were then band filtered in the physiologically relevant theta (4-7 Hz), alpha (8-12 Hz), and beta (13-30 Hz) frequency bands and band-specific epoched activity was source localized using the SBL algorithm.

SBL’s data-driven sparsity constraints reduce the effective number of sources considered at any given time as a solution, thereby reducing the uncertainty of the inverse solution. Thus, not only can higher channel density data yield source solutions, the ill-posed inverse problem can also be solved by imposing more aggressive constraints on the solution to converge on the source model at lower channel densities, as supported by prior research ^68,69^. We also benchmarked the SBL algorithm to show that it produces evidence-optimized inverse source models at 0.95AUC relative to the ground truth ^64,65^, and that the regions of interest (ROI) estimates resulting from this cortical source mapping have high test-retest reliability (Cronbach’s alpha=0.77, p<0.0001 ^26^).

For the source space activations, ROIs were based on the standard 68 brain region Desikan-Killiany atlas ^70^ using the Colin-27 head model ^71^. Artifacts remaining in source space were removed across all sessions and subjects using the >3 mad criterion. We compared the change in source ROI activity between pre- and post-sessions using rm-ANOVA with Greenhouse–Geisser significance correction applied. Spearman’s correlations were used to analyze neural change and cognitive performance change associations across sessions.

## 3. Results

### 3.1 Baseline Group Comparisons

**Table 1** shows demographics of both groups with regard to age, gender and ethnicity. There were no group differences in these demographics.

**Table 1.** Summary of participant demographics and baseline behaviors. Mean ± standard deviation data are shown for age in years, and gender and ethnicity are shown as percentage of total sample. Age was compared between groups using the non-parametric Wilcoxon Rank Sum test, and gender and ethnicity variables were compared between groups using Chi-square tests.

| Demographics | Intervention<br>(n=22) | Control<br>(n=21) | Group diff p-val |
| --- | --- | --- | --- |
| Age (years) | 28.27 ± 3.68 | 29.67 ± 4.96 | 0.407 |
| Gender n (%) |  |  | 0.700 |
| Male | 10 (45.45) | 12 (57.14) |  |
| Female | 10 (45.45) | 8 (38.10) |  |
| Ethnicity n (%) |  |  | 0.44 |
| Caucasian | 11 (50) | 9 (42.86) |  |
| Black/African American | 1 (4.55) | 1 (4.76) |  |
| Native Hawaiian or Other Pacific | 0 | 0 |  |
| Asian | 3 (13.64) | 8 (38.10) |  |
| Native American | 0 | 0 |  |
| More than one ethnicity | 5 (22.73) | 2 (9.52) |  |
| Other | 1 (4.55) | 1 (4.76) |  |

### 3.2 Behavioral and Cognitive Results

**Table 2** shows scores for the subjective scales in both groups and at both pre and post sessions. There were no baseline group differences. Notably, amongst burnout measures, only emotional exhaustion was high (greater than mid-scale score of 18). MBI depersonalization was low and personal accomplishment was high and these two subcomponents did not indicate burnout. Between-group rm-ANOVA comparisons with session as within-subject factor showed no main effects nor any interaction with sessions for any of the measures of mindfulness, well-being, self-compassion and burnout (all p>0.1).

**Table 2.** Summary of subjective outcome assessments from pre and post sessions (Intervention Group, n =22; Control Group, n = 21). Mean ± standard deviation data are shown for all variables. Data were checked for normality and pre vs. post within-group differences were compared using parametric/non-parametric tests. Mindfulness scores had a 6-point max. Well-being and self-compassion scores had a 5-point max. MBI: Maslach Burnout Inventory included EE: Emotional Exhaustion scores with 36 max, PA: Personal Accomplishment scores with 32 max and DP: Depersonalization scores with 20 max.

| Outcomes | Well-being Intervention | Control |
| --- | --- | --- |

**Table 2.** Summary of subjective outcome assessments from pre and post sessions (Intervention Group, n =22; Control Group, n = 21).
|  | Pre | Post | Pre | Post |
| --- | --- | --- | --- | --- |
| Mindfulness | 3.47 ± 0.66 | 3.34 ± 0.85 | 3.13 ± 0.75 | 3.31 ± 0.74 |
| Well-being | 3.25 ± 0.68 | 3.37 ± 0.62 | 3.32 ± 0.56 | 3.32 ± 0.66 |
| Self-Compassion | 2.86 ± 0.74 | 2.95 ± 0.70 | 3.08 ± 0.71 | 3.15 ± 0.66 |
| MBI EE | 26.95 ± 13.46 | 25.68 ± 12.21 | 24.05 ± 9.97 | 23.05 ± 12.15 |
| MBI PA | 31.52 ± 10.51 | 32.18 ± 10.08 | 32.86 ± 6.57 | 34.67 ± 7.58 |
| MBI DP | 5.67 ± 4.17 | 6.09 ± 4.16 | 9.29 ± 7.74 | 7.57 ± 7.19 |

**Table 3** shows cognitive performance outcomes in both groups at both pre and post sessions. Performance measures were compared using rm-ANOVAs; significant results were fdr-corrected for multiple comparisons. Processing speed on the selective attention task was the only measure with a significant group x session interaction (F_2,39_ = 12.71, p=0.005 fdr-corrected, bias corrected effect size (Hedges’ g) = 1.13); there were no main effects (p>0.3). Within-group post-hoc test showed that selective attention processing speed was improved at post-relative to pre-session for the intervention group (p = 0.019) and worsened for the control group (p = 0.022) (Figure 2).

**Figure 2.**
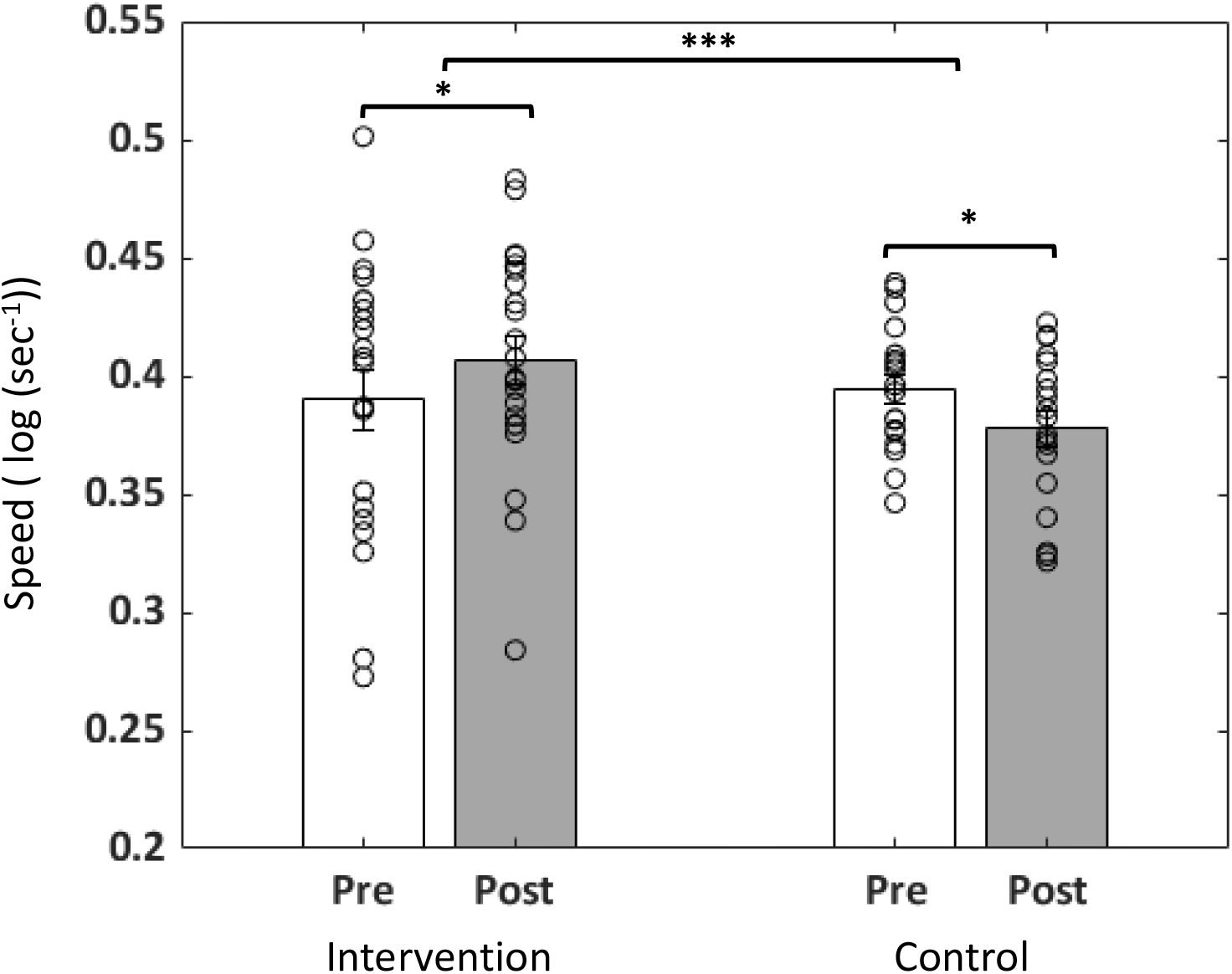
Changes in Selective Attention Processing Speed by session and group. Processing speed improved for the well-being intervention group and worsened for the control group. Individual data points are shown as scatterplot; bar length shows mean and error bars as standard error of mean *p<.05, ***p<.005

**Table 3.**
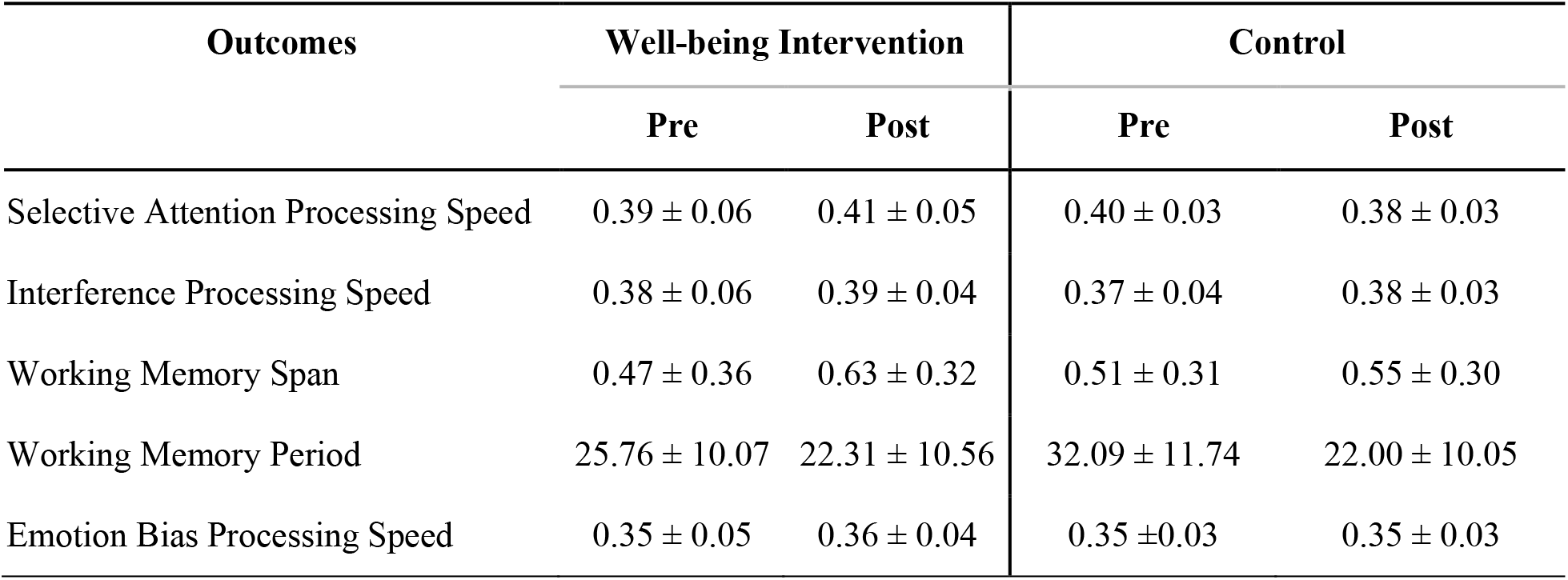
Summary of cognitive outcomes obtained in both groups. Processing speed units are log(sec^-1^) as the log inverse of the response time in seconds. Working memory (WM) span is out of 8 max item span and is normalized to 1. Length of the WM period is in seconds.

| Outcomes | Well-being Intervention |  | Control |  |
| --- | --- | --- | --- | --- |
|  | Pre | Post | Pre | Post |
| Selective Attention Processing Speed | 0.39 ± 0.06 | 0.41 ± 0.05 | 0.40 ± 0.03 | 0.38 ± 0.03 |
| Interference Processing Speed | 0.38 ± 0.06 | 0.39 ± 0.04 | 0.37 ± 0.04 | 0.38 ± 0.03 |
| Working Memory Span | 0.47 ± 0.36 | 0.63 ± 0.32 | 0.51 ± 0.31 | 0.55 ± 0.30 |
| Working Memory Period | 25.76 ± 10.07 | 22.31 ± 10.56 | 32.09 ± 11.74 | 22.00 ± 10.05 |
| Emotion Bias Processing Speed | 0.35 ± 0.05 | 0.36 ± 0.04 | 0.35 ± 0.03 | 0.35 ± 0.03 |

### 3.3 Neural Results

As the selective attention task showed intervention-related performance differences, i.e., processing speed improvement on target trials, we analyzed EEG data for this specific task. Figure 3A shows ERSP time-frequency plots for visual electrodes (O1 and O2) time-locked to selective attention target stimuli. Peak target post-stimulus processing occurred in the 100-300 ms time period across all sessions and subjects. Event-related synchronization (ERS) in the alpha frequency band (8-12 Hz), especially in the baseline pre-session that was reduced in the post-session (demarcated by red boxes in Figure 3A), dominated peak post-stimulus neural activity. We source-localized this visual alpha activity and quantified the magnitude of source activity in visual ROIs (Figure 3B). Visual ROIs included left and right visual striate pericalcarine ROIs and extra-striate lingual, lateral occipital, cuneus and fusiform ROIs. The magnitude of visual alpha source activity was reduced at the post relative to pre-session in both groups such that the session x group rm-ANOVA showed a main effect of session (F_1,38_ = 6.93, p=0.012) but no main effect of group (p>0.5) and no group x session interaction (p>0.9). However, the extent of post vs. pre visual alpha suppression was significantly correlated with the improvement in selective attention speed (partial Spearman correlation controlling for group, r = -0.32, p<0.05, Figure 3C). This correlation showed that individuals with greater increase in selective attention processing speed at the post-relative to pre-session showed greater visual source alpha suppression.

**Figure 3.**
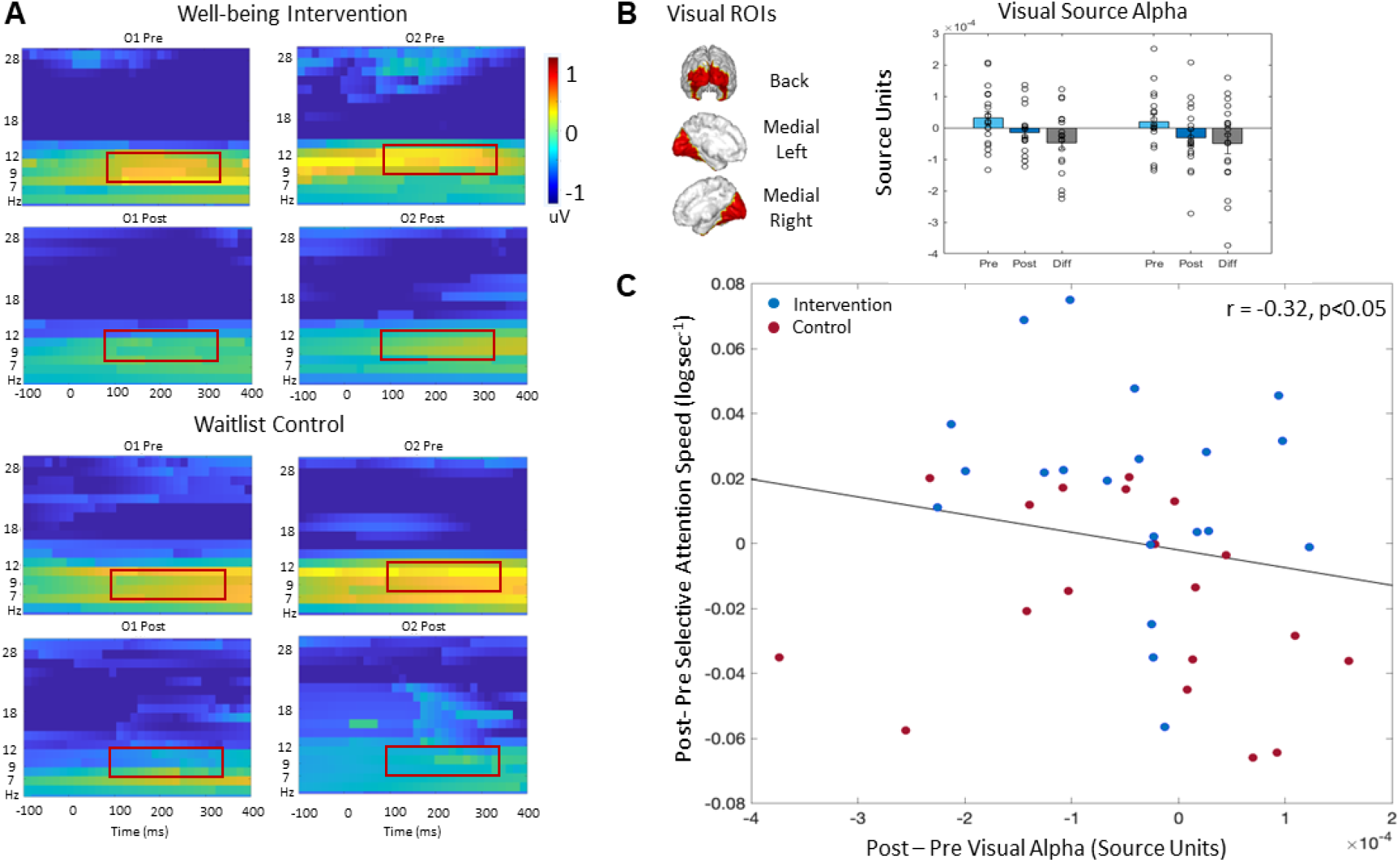
Neural Activity on the Selective Attention Task by session and group. (A) ERSP responses are shown in the well-being intervention group (top) and the control group (bottom) at occipital (O1:left and O2:right) electrodes at the pre- and post-sessions. Visual target event-related synchronization in the alpha band (8-12 Hz) is shown by red boxes. (B) Visual source ROIs are shown and the alpha activity in these ROIs is quantified at right. Individual data points are shown as scatterplot with bar length showing mean and error bars depicting standard error of mean. Both groups showed visual source alpha suppression at post-relative to pre-session. (C) Partial Spearman correlation controlling for group shows a significant inverse relationship between post minus pre change in selective attention processing speed vs. post minus pre change in visual alpha source activity.

### 3.4 Experience Feedback

These ratings were obtained at the end of the year by which time both groups had undergone the intervention. Average overall experience rating was very good to good (mean ± standard deviation, 3.70 ± 0.98) on a 5 point Likert scale with 5 being excellent. **Table 4** shows the group specific ratings with no significant between-group differences. Additionally, 57% of all participants said “yes” in response to “*As a result of this experience, do you expect your behavior to change in the future?*”. We extracted themes for qualitative responses on what they particularly liked/learned from the program and/or what could be improved. These themes included attention to lifestyle factors (such as sleep and exercise as determinants of well-being), learning mindfulness and compassion, and that the study generated a sense of community. Some participants voiced a need for structural change to address the issues faced by medical scientists Proportions of these themes in each group are shown in **Table 4**; there were no group differences.

**Table 4.**
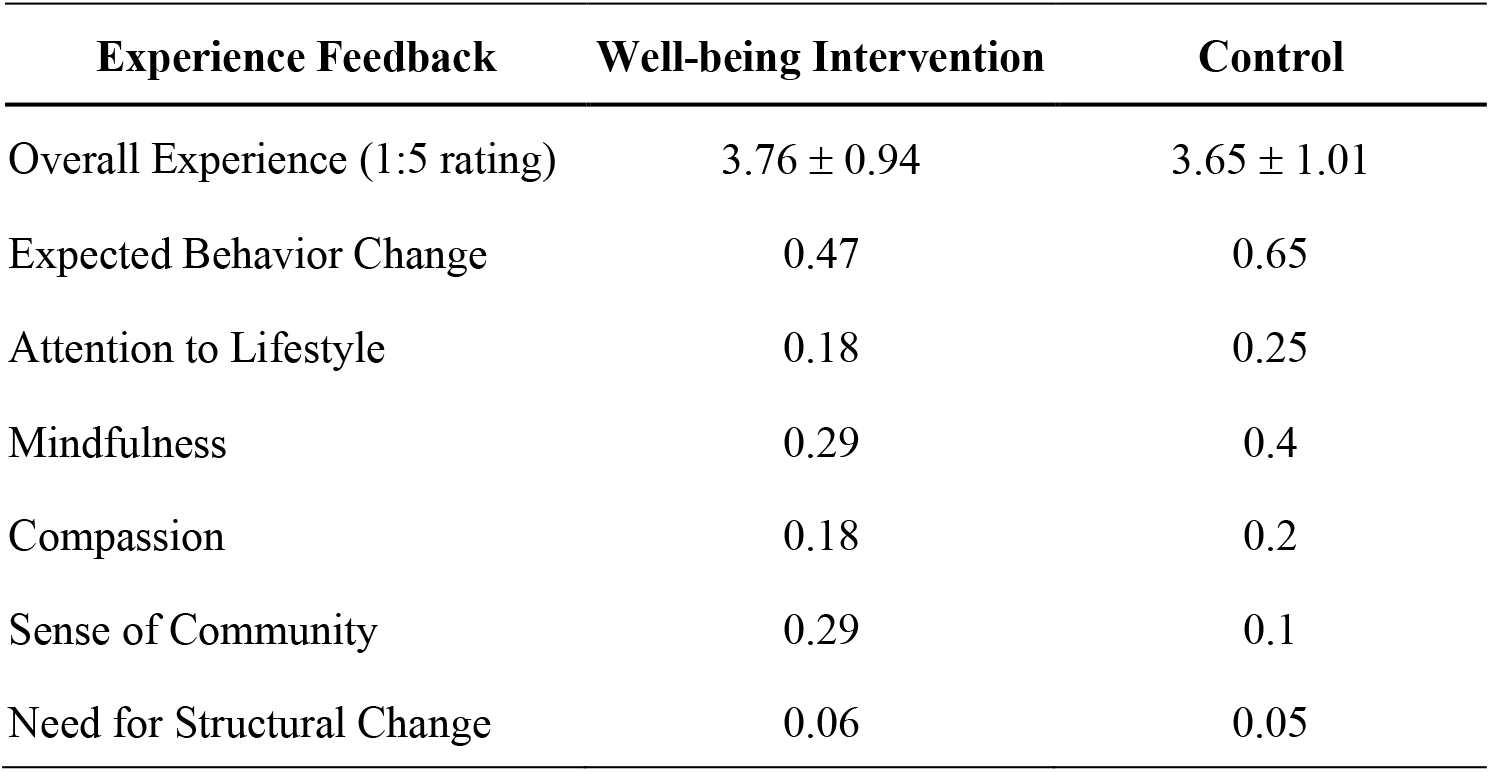
Summary of experience feedback outcomes obtained after both groups had undergone the intervention. Overall experience was rated on a 1-5 Likert Scale; expected behavior change as a result of the intervention had a yes/no response; the proportion of individuals providing a yes response are shown. The proportion of group members who brought up specific themes in the qualitative responses are also shown. There were no significant group differences in any feedback outcome variables.

| Experience Feedback | Well-being Intervention | Control |
| --- | --- | --- |
| Overall Experience (1:5 rating) | 3.76 ± 0.94 | 3.65 ± 1.01 |
| Expected Behavior Change | 0.47 | 0.65 |
| Attention to Lifestyle | 0.18 | 0.25 |
| Mindfulness | 0.29 | 0.4 |
| Compassion | 0.18 | 0.2 |
| Sense of Community | 0.29 | 0.1 |
| Need for Structural Change | 0.06 | 0.05 |

## 4. Discussion

This study investigated the outcomes of a brief quarter-long well-being intervention among M.D./Ph.D. trainees and graduates. Our goal was to enhance wellness in this cohort and address burnout. The program included three online group sessions with psychological health experts who provided mindfulness skills and led discussions on how to manage work-life challenges with wearable-based monitoring of lifestyle factors of sleep and exercise. We evaluated both subjective and objective outcomes of the intervention in a randomized controlled study design. Overall, the intervention was very feasible to deliver with all participants completing the intervention. Subjective outcomes of mindfulness, self-compassion, well-being and burnout showed no significant change but objective assessments demonstrated that selective attention processing speed was improved. EEG-based neural results suggested that the improvement in selective attention speed may be related to on-task visual alpha suppression – a neural marker for suppression of distractions ^72–76^.

While the well-being intervention indicated by full participation and completion, reasons for not observing significant changes on subjective measures could be due to intensity of the intervention or differences in style of the intervention compared to prior studies. For example, Krasner et al. (2009) implemented an 8-week intensive educational program (2.5 hours/week, 7-hour retreat) in mindfulness, communication, and self-awareness in primary care physicians. Pflugeisen et al., (2016) also implemented a mindfulness program in physicians over 8-weeks with weekly one-hour group teleconference coaching calls, weekly 5-7 minutes mindfulness training videos, and three 90-minutes in-person sessions. Our study had only three 90-minutes online group sessions. Yet, notably, the feedback surveys from both groups that underwent the intervention suggested an overall positive experience with emergent themes of learning mindfulness, compassion, sense of community and attention to lifestyle factors. Of all the participants, 57% reported that they would engage in future well-being behaviors as a result of the intervention, which suggests long-term benefit that can be evaluated by future follow-up assessments.

We measured objective cognitive outcomes in the domains of selective attention, interference processing, working memory and emotion bias. These assessments were adaptive and with a threshold to maintain accuracy at ∼80% so that ceiling effects would not occur with repeat administration. Cognitive outcomes showed a significant post-versus pre-intervention improvement in selective attention processing speed in the well-being intervention group, while the control group had a decrease at their repeat session. Such intervention-driven modulation of visual attention is consistent with prior studies of meditation-based psychosocial intervention ^77–79^. Digital meditation practice in young adults and children over a 6-8 week period can improve selective attention skills ^41,80^; in those studies, repeat test administration without intervention could be associated with worse performance, potentially due to boredom. Our prior digital meditation studies also found improvements in interference processing, which were potentially not observed here due to differences in intervention dose and style. Indeed, a review of cognitive effects of mindfulness trainings by Chiesa et al ^37^ concluded that cognitive functions show differential sensitivity to mindfulness-based interventions governed by varying factors in empirical studies, such as intervention study design, duration and participating populations.

In parallel with the change in selective attention speed, visual alpha oscillations were suppressed at the post-relative to pre-session. Alpha band oscillations are considered a sensory suppression mechanism in visual selective attention ^72–76^. Those studies have shown that sensory visual alpha suppression is controlled by top–down signals from frontoparietal attention networks. Thus, suppression of visual alpha band power following a stimulus is thought to measure attentional selection. Selective attention requires one to simultaneously attend to goal-relevant target stimuli while ignoring goal-irrelevant non-target stimuli. Alpha suppression for target stimuli (but not for non-targets) can facilitate selection. Post-stimulus visual processing usually also shows theta/alpha band ERS related to underlying spiking of visual sensory neurons ^81^, which is distinct from alpha suppression driven by top-down modulation. Here, we found that target stimuli had greater visual alpha suppression at the post-intervention session, suggesting improved attention selection. However, both groups at the group-average level showed this effect, which might be attributed to practice effects with repeat exposure in the control group. To investigate individual level effects, i.e., whether this plasticity in visual alpha may be relevant to change in selective attention performance speed, we performed a partial correlation controlling for group as a factor. We found a significant inverse correlation between improvement in selective attention processing speed and negative change in visual alpha. This result suggests that visual alpha suppression may underlie the individual-level improvement in cognitive performance. The relatively small effect size of the neural modulation suggests future studies need to replicate this outcome with larger sample sizes and using stronger intervention and control groups.

To the best of our knowledge, this is the first randomized, controlled study of a mindfulness-integrated intervention conducted with medical scientists and that assessed subjective and objective cognitive outcomes and neurophysiological measures. However, this study has limitations. Our intervention protocol, though highly feasible with online interactions, was brief and of low intensity compared to prior interventions conducted with physician cohorts ^18,20,22–24^. Prior work has also suggested that organization-directed workplace interventions could be effective for addressing burnout ^82^. The need for structural change was noted in the feedback survey by ∼5% of our participants. Hence, future studies may explore how mindfulness-integrated interventions can be deployed at a larger scale within an organizational framework that seeks to enhance physician well-being. It is also possible that our cohort of M.D./Ph.D. trainees and graduates did not detect results that differ for each group. Targeting a uniform cohort, such as early trainees who can build foundational mindfulness and well-being skills as part of their education, may lead to desirable long-term outcomes.

In conclusion, this randomized, controlled study sought to examine the effects of a mindfulness-integrated psychological intervention on improvement of subjective well-being related behaviors, cognitive functioning and associated electrophysiological dynamics. Despite the low intensity training, we found significant improvements in selective attention in the intervention group; this cognitive plasticity was correlated with alpha suppression in the visual cortex. Participants rated their intervention experience as very good; a majority plan to change their well-being related behaviors as a result of the intervention. Participants liked that the intervention focused on lifestyle factors, mindfulness and compassion, and that the study offered a sense of community. Based on these findings, we believe that a larger scale implementation of such an intervention integrated within the education curriculum may be useful for increasing well-being among physician scientist trainees and graduates. Overall, we posit that the well-being of our physicians and physician scientist workforce should be a pro-active and vital component of training to reduce burnout in this community and mitigate its deleterious consequences for the healthcare system.

## Data Availability

The dataset generated and analyzed during the current study will be available from a datadryad.org repository upon publication.

## Acknowledgements

This work was supported by a grant from the National Institutes of Health, NIH/NIGMS 1T32GM121318-01 (DMR, HHP), involved UCSD-MSTP trainees supported by NIH/NIGMS 5T32GM007198, and a Research Career Scientist Award from the Veterans Administration (BX005229 to HHP). We thank Alankar Misra for software development of the *BrainE* platform and UCSD students who assisted with data collection. The *BrainE* software is copyrighted for commercial use (Regents of the University of California Copyright #SD2018-816) and free for research and educational purposes.

## Author Contributions

SJ contributed to all data management, data analytics and wrote the first draft of the manuscript. JN managed and analyzed the data. SRP and JKM collected and managed the data. IG contributed to data analytics and manuscript writing and editing. DSR, DC, DTR, EB, PAI, NC, DMR, HHP and JM contributed to study conception and design and conduct of the study. JM supervised all data collection, data management, analytics, and manuscript writing. All authors contributed to manuscript edits, revisions, and approved the submitted version.

## Conflict of Interest

The authors declare no conflict of interest.

## Notes

### Competing Interest Statement

The authors have declared no competing interest.

### Clinical Trial

ISRCTN16736293

### Author Declarations

All participants provided written informed consent for study participation in accordance with the Declaration of Helsinki, and the UCSD Institutional Review Board approved all experimental procedures.

